# Lower Risk of Multisystem Inflammatory Syndrome in Children (MIS-C) with the Delta and Omicron variants of SARS-CoV-2

**DOI:** 10.1101/2022.03.13.22272267

**Authors:** Jonathan M Cohen, Michael J Carter, C Ronny Cheung, Shamez Ladhani, Evelina PIMS-TS Study Group

## Abstract

Little is known about the MIS-C risk with different SARS-CoV-2 variants. In Southeast England, MIS-C rates per confirmed SARS-CoV-2 infections in 0-16 years-olds were 56% lower (rate ratio, 0.34; 95%CI, 0.23-0.50) during pre-vaccine Delta, 66% lower (0.44; 0.28-0.69) during post-vaccine Delta and 95% lower (0.05; 0.02-0.10) during the Omicron period.

## Introduction

Multisystem Inflammatory Syndrome in Children (MIS-C, also known as PIMS-TS) is an acute inflammatory disorder, sharing features of Kawasaki disease and toxic-shock syndrome, and typically occurring 2-6 weeks after exposure to SARS-CoV-2, mainly in children and young people (CYP). First described in England in April 2020,^1^ this condition is now widely reported worldwide. Early estimates suggested a risk of MIS-C of 1 in 3-4,000 infected children.^2,3^ Whether this risk is sustained with new SARS-CoV-2 variants remains unknown. We compared daily cases of MIS-C against SARS-CoV-2 infection in children in the same geographical area of Southeast England to determine relative rates of MIS-C during the Alpha, Delta and Omicron waves over a 22-month period.

## Methods

### Data Sources

We utilised prospective data from the NHS South Thames Paediatric Network (STPN), which manages all MIS-C cases amongst 1·5 million children in South-East England, to assess trends over time. All patients aged 0–16 years with suspected MIS-C in the Network are discussed in a daily multi-disciplinary meeting, with diagnosis made according to RCPCH criteria, unchanged since September 2020.^4^

We compared MIS-C cases with two independent SARS-CoV-2 infection datasets. We used publicly available UK Health Security Agency (UKHSA) case numbers aggregated for age bands 0–4, 5–9 and 9–14 years across London and South East England, which records daily positive PCR and rapid-antigen tests from both healthcare and community testing within defined geographical regions.^5,6^ These were matched to STPN catchment, weighting cases to reflect child population distributions according to area population estimates from the Office for National Statistics (ONS).^7^ Since this dataset could potentially be biased by evolving changes in testing behaviour, we also compared MIS-C cases to community infection rates in 2-11 year olds, obtained from the ONS COVID-19 Infection Survey.^8^ This survey randomly selects individuals in private households for fortnightly PCR tests, deriving regional and national estimates of proportions infected in different age-groups irrespective of symptom status.

### Normalisation and Data Presentation

All three datasets were independently normalised to the peak of the Alpha variant wave in January 2021, allowing comparison with future waves, and plotted against time. MIS-C cases were smoothed over 7 days and plotted against a date corresponding to 40 days prior to their hospital admission, allowing for the lag from SARS-CoV-2 infection to hospitalisation. This interval was chosen as it provided the closest parallel in rise of SARS-CoV-2 infection and MIS-C cases during the Alpha variant wave.

### Definition of Variant Wave time periods

Using publicly available data on relative incidence of SARS-CoV-2 variants, we defined conservative time periods for each variant wave as the dates when the specific variant was responsible for ≥90% of typed cases in both the London and SE England regions.^9,10,11^ These correspond to 28 December 2020 to 7 March 2021 (Alpha), 13 June 2021 to 5 December 2021 (Delta) and 25 December 2021 to 1 Feb 2022 (Omicron). To control for the possible contribution of vaccination to changing MIS-C rates, an even more conservative period of the Delta variant wave was defined by restricting the time period until 12 September 2021 (week 36, 2021), when COVID-19 vaccination was first recommended for 12-15-year-olds in England.^12^

### Calculation of relative incidence of MIS-C

We calculated MIS-C incidence rates in two ways, using both the reported SARS-CoV-2 infection rates and community SARS-CoV-2 positivity rates as independent denominators. MIS-C cases were used as raw case numbers in the defined time periods as above. SARS-CoV-2 infection rates published by UKHSA and smoothed over 30 days were used as denominators to calculate MIS-C rates for each of the time periods above.

In the second analysis, the ONS COVID infection Survey provided estimates of percentage positivity at fortnightly time points. These values were imputed on an unchanged basis to the prior 13 days, and then summed over each of the defined time-periods to estimate infection rates, which were then used to calculate MIS-C rates. Using the Alpha period as baseline, we calculated relative rate ratios with 95% confidence intervals (CI) in MIS-C rates during the pre-vaccine Delta, post-vaccine Delta and the Omicron periods.

## Results

Compared with the Alpha wave, we found fewer MIS-C cases relative to SARS-CoV-2 infections during both pre-vaccine Delta, post-vaccine Delta and Omicron waves in CYP. Using confirmed SARS-CoV-2 infection cases as the denominator, MIS-C cases were 56% lower (rate ratio, 0.34; 95%CI, 0.23-0.50; P<0.001) during the pre-vaccine Delta period compared to the alpha period, 66% lower (0.44; 95%CI, 0.28-0.69; P<0.001) during post-vaccine Delta and 95% lower (0.05; 95%CI, 0.02-0.10; P<0.001) during the Omicron period compared to Alpha variant wave (1 in 432 cases during Alpha, 1 in 977 during pre-vaccine Delta, 1 in 1260 during post-vaccine Delta and 1 in 8315 during Omicron). Using community infection positivity rates in 2⍰11-year-olds from the ONS COVID Infection Survey as the denominator, the relative rates were 59%, 62% and 94% lower, respectively.

## Discussion

Testing for SARS-CoV-2 infection was not widely available during the first pandemic wave caused by the wild-type Wuhan strain in Spring 2020. Consequently, we did not include the first wave in this analysis and, instead, used the Alpha wave as a benchmark. Free, accessible community PCR testing became widely available in England since June 2020 and, from March 2021, secondary school-aged students (11⍰18 year-olds) were asked to undertake twice-weekly home testing using rapid-antigen tests and report their results online. Despite the increased testing practices, patterns of reported infections closely matched those of the ONS COVID-19 Infection Survey, which captured both symptomatic and asymptomatic infections in each age-group in the community every two weeks throughout the surveillance period. Reassuringly, MIS-C rates in children were very similar with both denominators, indicating true change in relative incidence of MIS-C during the different SARS-CoV-2 variant waves.

The lower MIS-C incidence during the pre-vaccine Delta wave was unlikely to be due to immunity from prior SARS-CoV-2 infection because nearly all Delta variant infections in children were primary infections and re-infections with Delta variant in children were uncommon during that period.^13^ Recent studies have shown that mRNA vaccination against COVID-19 helps protect against MIS-C.^14^ For this reason, we assessed MIS-C risk during the Delta wave before and after COVID-19 vaccination was recommended for 12-15 year-olds in England. The lower MIS-C rate in the pre-vaccine Delta period in comparison to the Alpha wave is, therefore, unlikely to be due to vaccination. Additionally, compared to many other countries, COVID-19 vaccine uptake among adolescents in England has been low, with only half of 12-15 years receiving the first dose by the end of 2021,^15^ while vaccination for 5–11-year-olds is planned for spring 2022. This likely explains the similar MIS-C rate reductions in the pre-vaccine and post-vaccine Delta periods compared to the Alpha wave. While prior infection and vaccination are unlikely to explain the lower MIS-C rates in the Delta wave, a more likely explanation may be viral mutations in key antigenic epitopes responsible for triggering the hyper-inflammatory response observed with MIS-C.^16^

The more recent Omicron wave, which first emerged in November 2021, was associated with very high cases numbers but we found an even lower relative MIS-C risk compared with previous variants. When the Omicron variant emerged in England more than half the primary school-aged children (up to 11 years) and nearly all secondary-school aged children (12-15) had SARS-CoV-2 antibodies consistent with prior infection and/or vaccination.^17^ Omicron possesses a number of new mutations in addition to those present in the Alpha, Beta and Delta variants, which allows the variant to at least partially evade both natural and vaccine-induced immunity, resulting in high rates of re-infections in previously-infected and vaccinated populations.^18^ Consequently, the relative contributions of host immunity and viral changes to the even lower MIS-C rates during the Omicron wave are obscured.

Both of the denominator data sets used have limitations. Changing testing behaviour as a bias of community infection rates is controlled for by using the ONS COVID Infection Survey data, which estimates infection rates by age group irrespective of symptom status and provides regionally- and nationally-representative community infection rates at fortnightly intervals. The finding of very similar rate reductions with the different variants using two independent denominators provides confidence that these findings are real and reflect true ongoing changes in incidence in the community which is consistent with our clinical experience on the frontline.

As more children become immune through natural SARS-CoV-2 infection and/or vaccination, and with the on-gong boosting of immunity following reinfections with the omicron variant, we predict that MIS-C will behave similarly to Kawasaki disease and become a sporadic condition occurring mainly in immunologically-naïve infants and toddlers.

## Data Availability

All data not publically available used in the present study are available upon reasonable request to the authors

## Acknowledgements

The authors are grateful to all the clinicians of the South Thames Paediatric Network and the Evelina London PIMS-TS Multidisciplinary Team for their contributions towards the networked care of children and young people with MIS-C.

## Author Contributions

JMC and MJC conceived the idea. Methodology was designed and formal analysis performed by JMC, and verified by MJC. Data interpretation was performed by JMC, MJC, SL and CRC. The manuscript was written by JMC, MJC, SL and CRC.

---

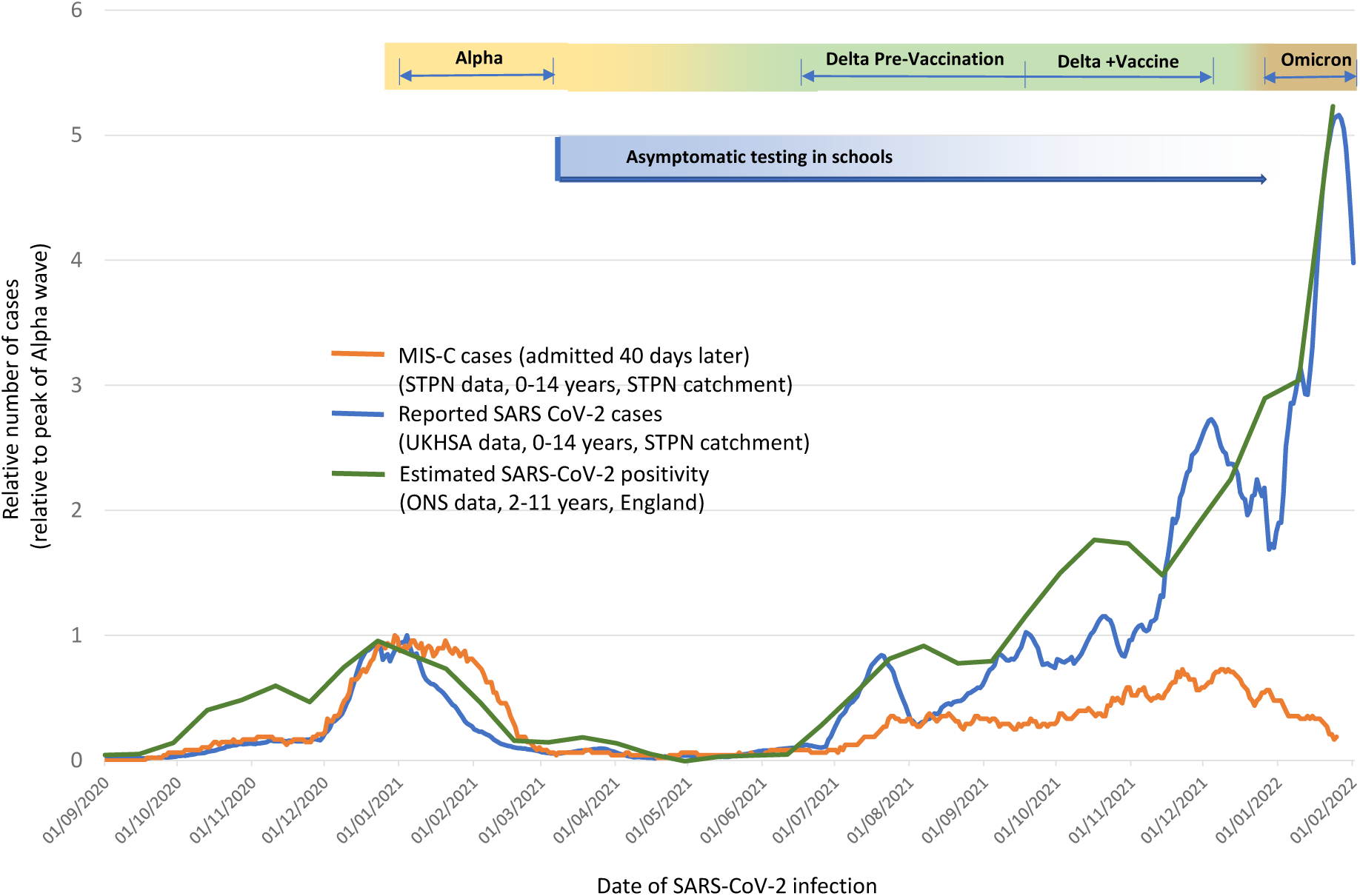

## References

1. Riphagen S, Gomez X, Gonzalez-Martinez C, Wilkinson N and Theokaris P. Hyperinflammatory shock in children during COVID-19 pandemic. Lancet, 2020; 395(10237):1607–1608.

2. Holm M, Hartling UB, Schmidt LS, et al. Multisystem inflammatory syndrome in children occurred in one of four thousand children with severe acute respiratory syndrome coronavirus 2. Acta Paediatr 2021; 110(9): 2581–3.

3. Payne AB, Gilani Z, Godfred-Cato S, et al. Incidence of Multisystem Inflammatory Syndrome in Children Among US Persons Infected With SARS-CoV-2. JAMA Netw Open 2021; 4(6): e2116420.

4. Royal College of Paediatrics and Child Heath. Guidance: Paediatric multisystem inflammatory syndrome temporally associated with COVID-19 2020. Available at: https://www.rcpch.ac.uk/sites/default/files/2020-05/COVID-19-Paediatric-multisystem-%20inflammatory%20syndrome-20200501.pdf. Accessed 24/02/2022.

5. UK Health Security Agency. Cases by specimen date age demographics [London Region]. Available at: https://coronavirus.data.gov.uk/details/cases?areaType=Region&areaName=London. Accessed 07/02/2022.

6. UK Health Security Agency. Cases by specimen date age demographics [South East Region]. Available at: https://coronavirus.data.gov.uk/details/cases?areaType=Region&areaName=SouthEast. Accessed 07/02/2022.

7. Office for National Statistics. Estimates of the population for the UK, England and Wales, Scotland and Northern Ireland (mid-2020). Available at: https://www.ons.gov.uk/peoplepopulationandcommunity/populationandmigration/populationestimates/datasets/populationestimatesforukenglandandwalesscotlandandnorthernireland. Accessed 08/02/2022).

8. Office for National Statistics. COVID Infection Survey. Available at: https://www.ons.gov.uk/peoplepopulationandcommunity/healthandsocialcare/conditionsanddiseases/datasets/coronaviruscovid19infectionsurveydata. Accessed 09/02/2022.

9. UK Health Security Agency. Variant of Concern Technical Briefing 5. Available at: https://assets.publishing.service.gov.uk/government/uploads/system/uploads/attachment_data/file/957631/Variant_of_Concern_VOC_202012_01_Technical_Briefing_5_Data_England.ods. Accessed 15/03/2022.

10. UK Health Security Agency. Variant of Concern Technical Briefing 23. Available at: https://assets.publishing.service.gov.uk/government/uploads/system/uploads/attachment_data/file/1018476/Variants_of_Concern_Technical_Briefing_23__Data_England.xlsx. Accessed 15/03/2022.

11. UK Health Security Agency. Variant of Concern Technical Briefing 38. Available at: https://assets.publishing.service.gov.uk/government/uploads/system/uploads/attachment_data/file/1060253/variants-of-concern-technical-briefing-38-data-england-11-March-2022.ods. Accessed 15/03/2022.

12. Powell A, Kirsebom F, Stowe J, et al. Adolescent vaccination with BNT162b2 (Comirnaty, Pfizer-BioNTech) vaccine and effectiveness against COVID-19: national test-negative case-control study, England. medRxiv 21267408 [Preprint]. December 10, 2021 [cited 21 March 2022]. Available from: https://doi.org/10.1101/2021.12.10.21267408.

13. Mensah A, Campbell C, Stowe J, et al. Risk of SARS-CoV-2 reinfections in children: prospective national surveillance, January 2020 to July 2021, England. medRxiv 21267372 [Preprint]. December 11, 2021 [cited 21 March 2022]. Available from: https://www.medrxiv.org/content/10.1101/2021.12.10.21267372.

14. Zambrano LD, Newhams MM, Olson SM, et al. Effectiveness of BNT162b2 (Pfizer-BioNTech) mRNA Vaccination Against Multisystem Inflammatory Syndrome in Children Among Persons Aged 12-18 Years -United States, July-December 2021. MMWR Morb Mortal Wkly Rep 2022;71:52-58. DOI: http://dx.doi.org/10.15585/mmwr.mm7102e1

15. UK Health Security Agency. Weekly Influenza and COVID-19 Surveillance Graphs 24/02/2022. Available at: https://assets.publishing.service.gov.uk/government/uploads/system/uploads/attachment_data/file/1057081/Weekly_COVID-19_and_Influenza_Surveillance_Graphs_w8.pdf. Accessed 01/03/2022.

16. Noval Rivas M, Porritt RA, Cheng MH, Bahar I, Arditi M. COVID-19-associated multisystem inflammatory syndrome in children (MIS-C): A novel disease that mimics toxic shock syndrome-the superantigen hypothesis. J Allergy Clin Immunol 2021. 147(1):57–59. doi: 10.1016/j.jaci.2020.10.008

17. Office for National Statistics. Coronavirus Infection Survey, antibody data, UK: 9 March 2022. Available at: https://www.ons.gov.uk/peoplepopulationandcommunity/healthandsocialcare/conditionsanddiseases/bulletins/coronaviruscovid19infectionsurveyantibodyandvaccinationdatafortheuk/9march2022. Accessed 21 March 2022.

18. Mallapaty S. COVID reinfections surge during Omicron onslaught. Nature 2022. Available at: https://www.nature.com/articles/d41586-022-00438-3. Accessed 24 March 2022.

